# A Digital-Ready Framework for Ongoing Performance Evaluation of India’s Integrated Disease Surveillance Program (IDSP) using Weekly Outbreak Reports: A Study from Bihar, 2016-2018

**DOI:** 10.64898/2026.08.13.26360363

**Authors:** Dhananjay Kumar Srivastava, S.D. Gupta, Nidhi Yadav

## Abstract

**Background:** Evaluation of public health surveillance systems is a programmatic obligation but has largely been conducted as a periodic, externally commissioned activity requiring dedicated resources and additional data collection. India’s Integrated Disease Surveillance Program (IDSP) generates continuous outbreak data through weekly reports but lacks a routine, embedded performance evaluation mechanism. This study assessed the quality of IDSP outbreak detection and response across multiple surveillance attributes and developed a weighted composite performance scoring framework using only routine program data.

**Methods:** A cross-sectional evaluation study was conducted across 38 districts of Bihar using secondary data from IDSP Central Surveillance Unit weekly outbreak reports for 2016 - 2018 (n=559 outbreaks). Six surveillance quality attributes were assessed - timeliness, completeness, representativeness, relative sensitivity, acceptability and flexibility. A weighted composite performance scoring scale was developed using expert opinion-derived attribute weightages (n=25 experts). District-level scores were computed and scaled to 100.

**Results:** Timeliness was the poorest-performing attribute, with fewer than 15% of outbreaks notified within 48 hours across all three years. Private sector participation was entirely absent - the acceptability score was 0 across all 38 districts for all three years. Completeness was the strongest attribute, exceeding 95% in all years. The mean composite score remained consistently low (23 - 27 out of 100) with widening inter-district disparity over time. Four districts (10.5%) scored 0 in all three years.

**Conclusions:** This study presents a dynamic, routine-data-based composite performance evaluation framework for IDSP outbreak detection and response. The modular, configurable framework functions at any administrative level (from block to national) and is compatible with digital health information platforms, enabling continuous, embedded performance monitoring without additional data collection. The framework has been registered as an Intellectual Property with the Government of India.

## BACKGROUND

CDC defined epidemiological surveillance as the ongoing systematic collection, analysis, and interpretation of health data, essential to the planning, implementation, and evaluation of public health practice, closely integrated with the timely dissemination of these data to those who need to know [1]. A functional surveillance system is an essential and crucial instrument for providing information for action [2]. Early detection of outbreaks is an important public health function of a surveillance system [3]. Integrated Disease Surveillance and Response (IDSR) and Integrated Disease Surveillance Program (IDSP) are intended to generate Early Warning Signals (EWS) of impending outbreaks and help initiate effective response in a timely manner [3, 4].

Surveillance has been largely interpreted and implemented as a component within vertical single disease control programs in most low- and middle-income countries including India [5]. Multiple un-coordinated single disease control programs overburden sub-national staff and result in duplication of efforts [6, 7] and the majority of them are heavily centralised [8, 9]. In 1998, the World Health Organization developed and advocated an Integrated Disease Surveillance and Response (IDSR) strategy [2, 10, 11, 12]. It aims to strengthen surveillance and response at each level of the health system, integrate existing multiple disease-specific surveillance systems, improve the use and flow of surveillance information, and link surveillance to public health actions [10, 11, 12]. India adopted the IDSR strategy in 2004, building on the National Surveillance Program for Communicable Diseases (NSPCD) (1997–2002) [11, 13]. IDSP is a district-centred integrated program of surveillance of communicable and non-communicable diseases [13]. It uses syndromic, presumptive and laboratory confirmation approaches to collect data through S, P and L reporting formats from identified reporting units on a weekly basis [13, 14]. The weekly outbreak (suspected) reports (narrative) compiled and published by the Central Surveillance Unit (CSU) contain information on outbreak detection, reporting source, date of onset, investigation and response for each outbreak reported from all districts and states.

### Need for multi-attribute evaluation

Public health surveillance systems should be evaluated periodically to ensure that problems of public health importance are being monitored efficiently and effectively [1]. Evaluation should include recommendations for improving quality, efficiency, and usefulness [1]. Several guidelines exist for evaluation of public health surveillance systems [1, 3, 15, 16, 17, 18, 19]. The CDC guidelines recommend assessment of ten attributes - simplicity, flexibility, data quality, acceptability, sensitivity, positive predictive value, representativeness, timeliness, stability and usefulness [1, 19]. The 2004 CDC framework for evaluating public health surveillance systems for early detection of outbreaks specifies timeliness, flexibility, sensitivity and positive predictive value as core capacity attributes [3].

A systematic review by Drewe et al. on evaluation of animal and public health surveillance systems revealed that quantitative approaches were applied more commonly compared to qualitative methods [20]. The most frequently assessed attributes were sensitivity, timeliness and data quality [20]. Calba et al. in a follow-up systematic review identified 49 distinct evaluation attributes grouped into four categories - effectiveness attributes (timeliness, sensitivity, representativeness, specificity, positive predictive value, completeness and reliability), functional attributes (acceptability, flexibility, data quality, stability, simplicity and portability), value attributes (usefulness, cost, efficacy, efficiency and impact) and organisational attributes (communication, data management and laboratory management) [21]. Assessment of attributes from a single category does not characterise the performance of a surveillance system across all dimensions identified in this framework [21]. Considerable variation in the approaches used for evaluation of outbreak detection methods in public health surveillance data has been noted, with no single approach of choice identified [22].

### Evaluation of IDSP - existing evidence and gaps

All evaluation studies of IDSP identified in the literature were limited to the district, block or state level and focused on specific diseases or selected individual attributes [23, 24, 25, 26, 27, 28, 29]. The attributes most commonly assessed in these studies were timeliness and completeness [30, 26, 31]. These studies were conducted either through externally commissioned assessments or by independent researchers. A routine, systematic mechanism for ongoing performance evaluation of IDSP outbreak detection and response does not exist within the program. The Joint Monitoring Mission (JMM) 2015, recommended developing a self-assessment tool for IDSP to monitor and assess early threats detected through event-based surveillance [32]. It further recommended development of a Monitoring and Evaluation mechanism for IDSP to foster the principle of information for action at state and national level [32].

### Need for a composite performance scoring framework

Scott et al. developed a conceptual framework for public health surveillance and action categorised into core and support activities measured with indicators [33]. This approach permits cost analysis, highlights areas amenable to integration and provides a focused approach to improvement [33]. Cavallaro et al. developed indicator scores by adopting quantitative criteria and targets from published surveillance guidelines, with components aggregated into composite measures [34]. Vivek Singh et al. developed a composite score from key surveillance quality indicators for assessment of IHR core capacity requirements in India [30]. Dashboard monitoring on the basis of a composite performance score enables identification of gaps in a surveillance system and facilitates timely mid-course correction [30]. Individual attribute scores assessed separately do not provide a single comparable and trackable measure of overall system performance across districts and over time. A composite performance score integrating multiple surveillance attributes with expert opinion-derived weightages addresses this limitation, providing a standardised framework applicable across districts and states. Very limited attempts have been made at developing composite scoring for disease surveillance systems [34, 30]. A composite scoring framework for IDSP outbreak detection and response has not been developed.

### Routine data and digital surveillance readiness

The weekly outbreak reports generated by the CSU of IDSP are available in the public domain and provide a continuous source of program data on outbreak detection and response across all districts and states. Use of these routine program data for performance evaluation eliminates the need for additional data collection. It makes systematic evaluation feasible at scale and on a continuous basis, independent of external commissioning or researcher-driven access to program data. India’s Vision 2035: Public Health Surveillance identifies transition to a responsive and predictive surveillance system through enhanced use of analytics, health informatics and data science, and improved real-time data-sharing mechanisms between centre and states, as the building blocks for the national public health surveillance system [35]. A composite performance scoring framework structured on routine surveillance data is compatible with digital health information platforms and can enable continuous, embedded performance monitoring within the surveillance system in alignment with this national vision.

This study aimed to assess the quality of IDSP outbreak detection and response in Bihar during 2016– 2018 across six surveillance performance attributes (timeliness, completeness, representativeness, relative sensitivity, acceptability and flexibility) and to develop a composite performance evaluation scoring scale for ongoing, program-embedded performance monitoring of outbreak detection and response at any administrative level.

## METHODS

### Study design and setting

A cross-sectional evaluation study was conducted to develop and demonstrate a composite performance scoring framework for IDSP outbreak detection and response using routine surveillance data. Bihar, India was selected as the demonstration setting for application of this framework. Bihar is the 3rd most populous state of India with a population of approximately 104 million (Census 2011). It has the lowest proportion of urban population (11.3%) among major Indian states and comprises 9 administrative divisions and 38 districts, with significant variation in health infrastructure and disease burden across districts.

### Data source

This study was based mainly on secondary data. The weekly outbreak reports generated by the CSU of IDSP, available in the public domain at www.idsp.nic.in were downloaded, compiled and analysed for the years 2016, 2017 and 2018. These reports are published weekly and contain structured information for each suspected outbreak - including date of onset, district and block, suspected disease, number of cases and deaths, age group affected, source of report (IDSP reporting unit, informer, media or community) and response action taken. A total of 559 suspected outbreaks were available for analysis: 246 in 2016, 142 in 2017 and 171 in 2018. Data were extracted into Microsoft Excel spreadsheets for cleaning and analysis.

### Surveillance quality attributes and indicator framework

Six surveillance quality attributes were selected for assessment: timeliness, completeness, representativeness, relative sensitivity, acceptability and flexibility. Indicators for each attribute were developed drawing upon the following established international frameworks: the ECDC Technical Document for evaluating public health surveillance systems [15]; the WHO protocol for assessing national communicable disease surveillance and response systems [16]; the WHO guide to assessing disease surveillance and response systems [17]; the CDC 2001 updated guidelines for evaluating public health surveillance systems [1]; the CDC 2004 framework for evaluating public health surveillance systems for early detection of outbreaks [3]; and the WHO framework for evaluating communicable disease surveillance systems [18].

Each attribute was scored on a Likert scale of 0–5, based on the proportion of outbreaks meeting a defined criterion. The indicator scoring framework is designed to be bidirectional - the scoring direction for each indicator can be set as positive or negative depending on programmatic expectations and context. The acceptability indicator illustrates this: private sector involvement is scored positively here, where increasing private sector participation is a program objective; in a setting where private sector already dominates and public sector reporting needs strengthening, the scoring direction can be reversed. The complete indicator framework and scoring criteria are presented in Table 1.

**Table 1:** Surveillance Quality Attributes, Indicators, Scoring Framework and Expert Opinion Weightage.

| Attribute | Indicator | Scoring (0–5) | Opinion Based Weightage (Median)* |
| --- | --- | --- | --- |
| **Timeliness** | Proportion of outbreaks notified within 48 hours of onset | 0%=0; 1–20%=1;<br>21–40%=2; 41–60%=3; 61–80%=4;<br>>80%=5 | 5 |
| **Representativeness** | Proportion of blocks (district-wise) reporting an outbreak in a year [Assumption: blocks not reporting does not mean no outbreak occurred] | 0%=0; 1–20%=1;<br>21–40%=2; 41–60%=3; 61–80%=4;<br>>80%=5 | 4 |
| **Relative Sensitivity** | District reporting the highest number of outbreaks per million population taken as reference; relative sensitivity of other districts computed against this | 0%=0; 1–20%=1;<br>21–40%=2; 41–60%=3; 61–80%=4;<br>>80%=5 | 4 |
| **Acceptability** | Proportion of outbreaks reported by private healthcare providers in a district [Scoring direction configurable: scored positively here where private sector participation is a program objective; can be reversed where private sector dominance is a concern] | 0%=0; 1–20%=1;<br>21–40%=2; 41–60%=3; 61–80%=4;<br>>80%=5 | 5 |
| **Completeness** | Proportion of outbreaks reported with Time (date of onset), Place (village/location) and Person (cases, age group, gender) | 0%=0; 1–20%=1;<br>21–40%=2; 41–60%=3; 61–80%=4;<br>>80%=5 | 5 |
| **Flexibility** | Proportion of outbreaks reported by IDSP that are not on the standard IDSP surveillance disease list | 0%=0; 1–20%=1;<br>21–40%=2; 41–60%=3; 61–80%=4;<br>>80%=5 | 4 |

\* Weightage factors derived from expert opinion survey (n=25). Each attribute score multiplied by weightage factor to compute weighted score.

**Table 2:** Details of Expert Group and Attribute-wise Weightage Scores Derived from Expert Opinion Survey.

| **Gender** | **Highest educational Qualification** | **Years of Experience** | **Timeliness** | **Representativeness** | **Relative Sensitivity** | **Acceptability** | **Completeness** | **Flexibility** |
| --- | --- | --- | --- | --- | --- | --- | --- | --- |
| M | MD | 21 | 5 | 4 | 4 | 4 | 4 | 4 |
| M | MBBS | 20 | 5 | 5 | 1 | 5 | 5 | 1 |
| M | PhD | 10 | 5 | 3 | 5 | 4 | 5 | 4 |
| M | MPH | 5 | 5 | 3 | 5 | 5 | 5 | 5 |
| M | MBBS | 8 | 5 | 5 | 4 | 5 | 5 | 4 |
| M | MD | 21 | 5 | 4 | 5 | 5 | 5 | 4 |
| M | MD | 5 | 5 | 4 | 5 | 5 | 4 | 3 |
| F | MD | 4 | 5 | 4 | 2 | 5 | 5 | 5 |
| M | MPH | 5 | 5 | 5 | 5 | 5 | 5 | 5 |
| F | MPH | 3.5 | 5 | 2 | 1 | 4 | 5 | 5 |
| F | MPH | 1 | 3 | 2 | 3 | 3 | 2 | 3 |
| M | MD | 7 | 5 | 5 | 4 | 5 | 5 | 4 |
| M | MD | 34 | 5 | 3 | 5 | 4 | 5 | 4 |
| F | MD | 6 | 5 | 5 | 5 | 5 | 5 | 5 |
| F | MD | 15 | 2 | 2 | 3 | 1 | 3 | 1 |
| M | MD | 7 | 5 | 4 | 2 | 5 | 5 | 1 |
| M | MPH | 10 | 5 | 5 | 5 | 5 | 4 | 5 |
| M | MBBS | 11 | 5 | 5 | 5 | 5 | 5 | 5 |
| M | Others | 15 | 2 | 3 | 3 | 3 | 3 | 4 |
| M | MBBS | 3 | 5 | 3 | 1 | 4 | 5 | 2 |
| M | PhD | 12 | 3 | 4 | 3 | 1 | 4 | 1 |
| M | MPH | 12 | 4 | 4 | 2 | 5 | 5 | 4 |
| F | PhD | 5 | 5 | 4 | 5 | 5 | 5 | 5 |
| M | MBBS | 0 | 4 | 4 | 4 | 4 | 4 | 4 |
| M | MD | 32 | 5 | 5 | 5 | 5 | 5 | 3 |
| **Median score** |  |  | **5** | **4** | **4** | **5** | **5** | **4** |

### Expert opinion and weightage derivation

The importance of each of the six surveillance attributes for outbreak detection and response was rated by 25 public health experts through a self-administered online questionnaire. Expert identification was based on published research in epidemiology and disease surveillance, supplemented by known program experts working in area of surveillance. Participating experts included 19 males and 6 females with qualifications ranging from MBBS to PhD and experience in epidemiology or disease surveillance ranging from less than one year to 34 years. Each expert rated the importance of each attribute independently on a scale of 1 (least important) to 5 (most important). This was a single-round expert opinion survey and not a formal Delphi process. The median score for each attribute across all expert responses was calculated and used as the weightage factor for that attribute.

### Composite performance score computation

For each district, the raw attribute score (0–5) was multiplied by the expert-derived weightage factor to produce a weighted score for that attribute. The sum of weighted scores across all six attributes gave a raw composite score. The maximum possible raw composite score was 135. This was scaled proportionally to a final score out of 100 to facilitate comparison across districts and tracking over time. The computation was performed independently for each district for each of the three years (2016, 2017 and 2018).

### Framework design and adaptability

The framework presented in this study is a proof-of-concept demonstration. The indicators, cut-offs and weightage factors are designed to be modular and adaptable. Indicators can be added as data availability improves or removed where data are not captured by routine program records. Scoring cut-offs can be adjusted to reflect the maturity of the surveillance system - a program in early stages may set an initial benchmark and progressively raise it as performance improves. Indicator scoring direction is configurable as described above. Expert-derived weightages can be re-derived using a locally constituted expert panel to reflect current program priorities.

The framework was demonstrated at district level in this study but is designed to function at any administrative level where routine surveillance data are available. It can be configured at block level, enabling district managers to grade the performance of their own administrative blocks and obtain granular information for targeted intervention. The same framework scales upward to state and national level without modification. Routine program data are the sole input, eliminating the need for additional data collection. Once configured within a digital health information platform, the framework can function on a continuous, near-real-time basis as outbreak data are entered - making performance evaluation an ongoing, embedded function of the surveillance system rather than a periodic external exercise.

### Intellectual Property

The composite performance evaluation scoring framework developed in this study has been registered as an Intellectual Property with the Copyright Office, Government of India [IP Registration Number: L-151075; Registration Date: 15 July 2024].

### Ethics and permissions

Ethical clearance was obtained from the Institutional Ethics Committee of IIHMR University, Jaipur, India. The study was based mainly on secondary data available in the public domain (www.idsp.nic.in). Permission was obtained from the concerned authority of IDSP Bihar for academic use of program data and for development of a performance monitoring framework. Data confidentiality was maintained throughout. Informed consent was obtained from all participants in the expert opinion survey prior to their participation.

## RESULTS

A total of 559 suspected outbreaks were reported from the 38 districts of Bihar during the study period - 246 in 2016, 142 in 2017 and 171 in 2018. The epidemiological profile of these outbreaks, including disease-wise distribution, age group, deaths and seasonal trends, has been reported separately as a companion paper.

District-wise attribute scores and composite performance scores are presented in color-coded tables to facilitate visual interpretation of performance patterns across districts and years. Five categories are applied consistently across all tables - grey indicates no outbreak was reported by the district during that year; red (1–25) indicates poor performance; light red (25–50) below average performance; yellow (50–75) above average performance; and green (75–100) good performance. The same color categories are applied in the geographic visualisation of composite scores presented in Figure 1. These categories are configurable - program managers may define two categories (below and above a defined threshold), four, or any number appropriate to program context and decision-making needs.

**Figure 1:**
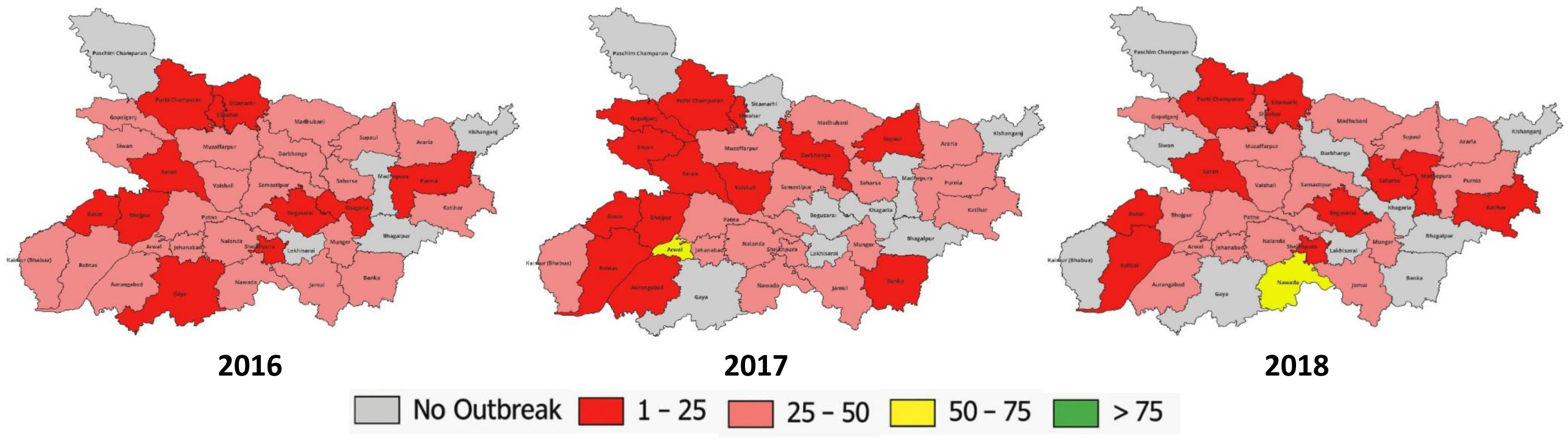
Geographic distribution of composite performance evaluation scores across districts of Bihar: 2016, 2017 and 2018. Color categories: Grey = 0 (no outbreak reported); Red = 1–25 (poor performance); Light Red = 25–50 (below average); Yellow = 50–75 (above average); Green = 75– 100 (good performance). Performance categories are configurable - program managers may define two categories (below and above a threshold), four as demonstrated here, or any number appropriate to program context and decision-making needs.

### Timeliness

The proportion of outbreaks notified within 48 hours of onset was 4.9% (12 of 246) in 2016, 12.0% (17 of 142) in 2017 and 5.8% (10 of 171) in 2018. District-wise timeliness scores are presented in Table 3. In 2016, Aurangabad achieved the highest timeliness score (3 of 5); 28 (73.7%) of 38 districts scored 0. In 2017, Kaimur scored 5 of 5; 29 (76.3%) districts scored 0. In 2018, Araria, Arwal and Aurangabad each scored 5 of 5; 29 (76.3%) districts scored 0. Consistent timeliness across all three years was not demonstrated by any district.

**Table 3:** Year-wise Comparison of Timeliness Scores across Districts of Bihar, 2016–2018.

| District | 2016 |  |  |  | 2017 |  |  |  | 2018 |  |  |  |
| --- | --- | --- | --- | --- | --- | --- | --- | --- | --- | --- | --- | --- |
|  | Total outbreak | <48 hours | % | Score | Total outbreaks | <48 hours | % | Score | Total outbreaks | <48 hours | % | Score |
| Araria | 3 | 1 | 33.3 | 2 | 2 | 0 | 0.0 | 0 | 1 | 1 | 100.0 | 5 |
| Arwal | 2 | 0 | 0.0 | 0 | 5 | 1 | 20.0 | 2 | 1 | 1 | 100.0 | 5 |
| Aurangabad | 2 | 1 | 50.0 | 3 | 3 | 0 | 0.0 | 0 | 1 | 1 | 100.0 | 5 |
| Banka | 9 | 0 | 0.0 | 0 | 1 | 0 | 0.0 | 0 | 0 | 0 | 0.0 | 0 |
| Begusarai | 2 | 0 | 0.0 | 0 | 0 | 0 | 0 | 0 | 1 | 0 | 0.0 | 0 |
| Kaimur | 8 | 3 | 37.5 | 2 | 4 | 4 | 100.0 | 5 | 0 | 0 | 0.0 | 0 |
| Bhagalpur | 0 | 0 | 0.0 | 0 | 0 | 0 | 0.0 | 0 | 0 | 0 | 0.0 | 0 |
| Bhojpur | 3 | 0 | 0.0 | 0 | 1 | 0 | 0.0 | 0 | 5 | 1 | 20.0 | 2 |
| Buxar | 3 | 0 | 0.0 | 0 | 2 | 0 | 0.0 | 0 | 1 | 0 | 0.0 | 0 |
| Darbhanga | 5 | 1 | 20.0 | 2 | 3 | 0 | 0.0 | 0 | 0 | 0 | 0.0 | 0 |
| East Champaran | 0 | 0 | 0.0 | 0 | 0 | 0 | 0.0 | 0 | 0 | 0 | 0.0 | 0 |
| Gaya | 5 | 0 | 0.0 | 0 | 0 | 0 | 0.0 | 0 | 0 | 0 | 0.0 | 0 |
| Gopalganj | 13 | 1 | 7.7 | 1 | 1 | 0 | 0.0 | 0 | 6 | 0 | 0.0 | 0 |
| Jamui | 17 | 1 | 5.9 | 1 | 12 | 0 | 0.0 | 0 | 3 | 0 | 0.0 | 0 |
| Jehanabad | 3 | 0 | 0.0 | 0 | 2 | 0 | 0.0 | 0 | 5 | 0 | 0.0 | 0 |
| Katihar | 10 | 0 | 0.0 | 0 | 7 | 3 | 42.9 | 3 | 1 | 0 | 0.0 | 0 |
| Khagaria | 1 | 0 | 0.0 | 0 | 0 | 0 | 0.0 | 0 | 0 | 0 | 0.0 | 0 |
| Kishanganj | 0 | 0 | 0.0 | 0 | 0 | 0 | 0.0 | 0 | 0 | 0 | 0.0 | 0 |
| Lakhisarai | 0 | 0 | 0.0 | 0 | 0 | 0 | 0.0 | 0 | 0 | 0 | 0.0 | 0 |
| Madhepura | 0 | 0 | 0.0 | 0 | 0 | 0 | 0.0 | 0 | 2 | 0 | 0.0 | 0 |
| Madhubani | 25 | 0 | 0.0 | 0 | 15 | 1 | 6.7 | 1 | 22 | 0 | 0.0 | 0 |
| Munger | 13 | 0 | 0.0 | 0 | 2 | 0 | 0.0 | 0 | 4 | 0 | 0.0 | 0 |
| Muzaffarpur | 14 | 0 | 0.0 | 0 | 16 | 0 | 0.0 | 0 | 29 | 2 | 6.9 | 1 |
| Nalanda | 12 | 0 | 0.0 | 0 | 14 | 0 | 0.0 | 0 | 10 | 1 | 10.0 | 1 |
| Nawada | 11 | 0 | 0.0 | 0 | 15 | 2 | 13.3 | 1 | 33 | 1 | 3.0 | 1 |
| Patna | 28 | 0 | 0.0 | 0 | 11 | 3 | 27.3 | 2 | 5 | 0 | 0.0 | 0 |
| Purnia | 2 | 0 | 0.0 | 0 | 2 | 1 | 50.0 | 3 | 6 | 0 | 0.0 | 0 |
| Rohtas | 15 | 1 | 6.7 | 1 | 3 | 0 | 0.0 | 0 | 1 | 0 | 0.0 | 0 |
| Saharsa | 4 | 0 | 0.0 | 0 | 4 | 0 | 0.0 | 0 | 1 | 0 | 0.0 | 0 |
| Samastipur | 8 | 0 | 0.0 | 0 | 3 | 1 | 33.3 | 2 | 6 | 0 | 0.0 | 0 |
| Saran | 2 | 0 | 0.0 | 0 | 1 | 0 | 0.0 | 0 | 2 | 0 | 0.0 | 0 |
| Sheikhpura | 1 | 0 | 0.0 | 0 | 4 | 1 | 25.0 | 2 | 1 | 0 | 0.0 | 0 |
| Sheohar | 1 | 0 | 0.0 | 0 | 1 | 0 | 0.0 | 0 | 2 | 0 | 0.0 | 0 |
| Sitamarhi | 3 | 0 | 0.0 | 0 | 0 | 0 | 0.0 | 0 | 1 | 0 | 0.0 | 0 |
| Siwan | 3 | 1 | 33.3 | 2 | 0 | 0 | 0.0 | 0 | 0 | 0 | 0.0 | 0 |
| Supaul | 5 | 1 | 20.0 | 2 | 2 | 0 | 0.0 | 0 | 6 | 1 | 16.7 | 1 |
| Vaishali | 11 | 1 | 9.1 | 1 | 4 | 0 | 0.0 | 0 | 8 | 1 | 12.5 | 1 |
| West Champaran | 2 | 0 | 0.0 | 0 | 2 | 0 | 0.0 | 0 | 7 | 0 | 0.0 | 0 |
| **Total** | **246** | **12** | **4.9** |  | **142** | **17** | **12.0** |  | **171** | **10** | **5.8** |  |

### Representativeness

Representativeness was measured as the proportion of blocks within each district that reported at least one outbreak in a year. The overall proportion of blocks reporting outbreaks across Bihar was 26.2% in 2016, 17.6% in 2017 and 16.7% in 2018. District-wise representativeness scores are presented in Table 4. In 2016, Munger achieved the highest score (5 of 5, with 100% of blocks reporting). In 2017, Arwal and Jamui each scored 4 of 5. In 2018, Nawada and Muzaffarpur each scored 5 of 5. The number of districts not meeting the representativeness criterion was 5 (13.2%) in 2016, increasing to 10 (26.3%) in both 2017 and 2018.

**Table 4:**
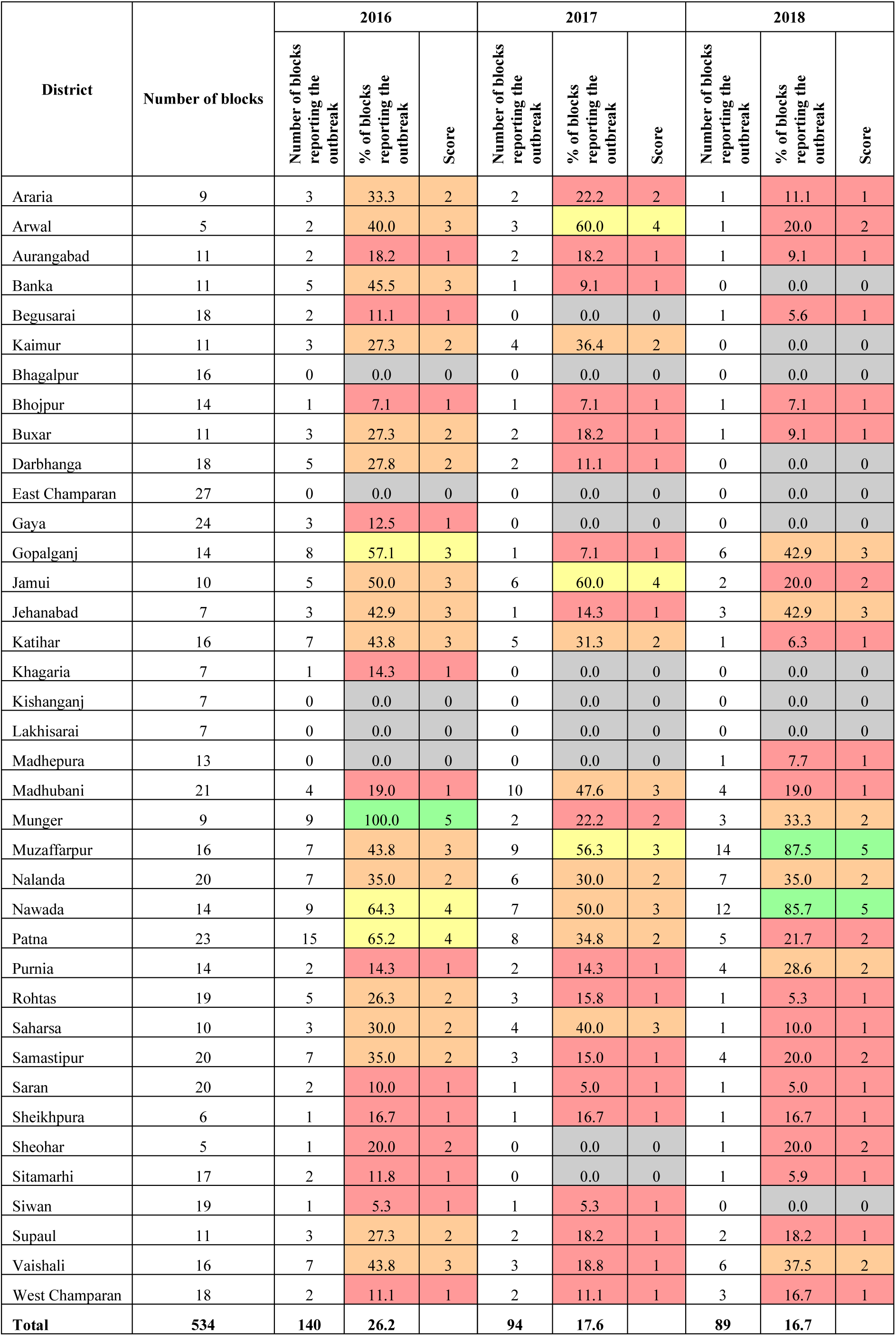
Year-wise Comparison of Representativeness Scores across Districts of Bihar, 2016–2018.

### Relative Sensitivity

The district reporting the highest number of outbreaks per million population in each year was used as the reference for calculating relative sensitivity of other districts. The reference districts were Munger in 2016 (8.59 outbreaks per million), Arwal in 2017 (6.37 per million) and Nawada in 2018 (12.67 per million). District-wise relative sensitivity scores are presented in Table 5. In 2016, Jamui and Munger each scored 5 of 5. In 2017, Arwal, Jamui, Nawada and Sheikhpura each scored 5 of 5. In 2018, only Nawada scored 5 of 5, followed by Muzaffarpur with a score of 3 of 5. The number of districts not meeting any relative sensitivity criterion was 5 (13.2%) in 2016 and 10 (26.3%) in both 2017 and 2018.

**Table 5:**
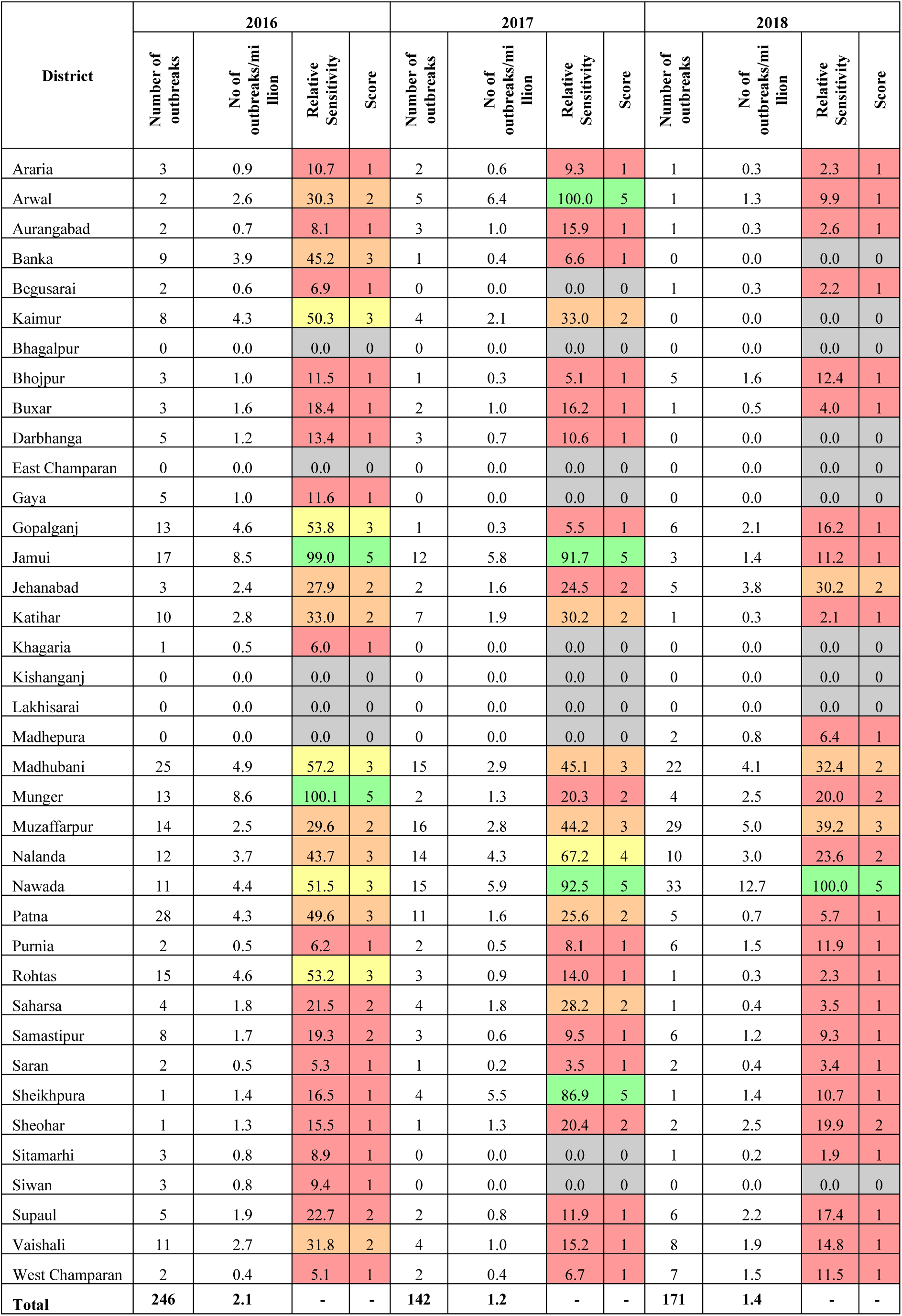
Year-wise Comparison of Relative Sensitivity Scores across Districts of Bihar, 2016–2018.

### Acceptability

Acceptability was measured as the proportion of outbreaks reported by private healthcare providers. Private providers did not report a single outbreak in any district during 2016, 2017 or 2018. The acceptability score was 0 for all 38 districts across all three years.

### Completeness

Completeness was scored on the proportion of outbreaks reported with Time (date of onset), Place (village or geographic location) and Person (number affected and age group). The overall completeness across Bihar was 95.0% (233 of 246 outbreaks) in 2016, 98.0% (139 of 142) in 2017 and 99.4% (170 of 171) in 2018. District-wise completeness scores are presented in Table 6. In 2016, 27 (71.1%) of 38 districts scored 5 of 5; 6 (15.8%) districts scored 0. In 2017, 27 (71.1%) districts scored 5 of 5; 11 (28.9%) districts scored 0. In 2018, 28 (73.7%) districts scored 5 of 5; 10 (26.3%) districts scored 0. Completeness was the highest-performing attribute across all three years.

**Table 6:** Year-wise Comparison of Completeness Scores across Districts of Bihar, 2016–2018.

| District | 2016 |  |  |  | 2017 |  |  |  | 2018 |  |  |  |
| --- | --- | --- | --- | --- | --- | --- | --- | --- | --- | --- | --- | --- |
|  | Total outbreaks | TPP reported in | Proportion of outbreaks with TPP | Score | Total outbreaks | TPP reported in | Proportion of outbreaks with TPP | Score | Total outbreaks | TPP reported in | Proportion of outbreaks with TPP | Score |
| Araria | 3 | 2 | 67 | 4 | 2 | 2 | 100 | 5 | 1 | 1 | 100 | 5 |
| Arwal | 2 | 1 | 50 | 3 | 5 | 5 | 100 | 5 | 1 | 1 | 100 | 5 |
| Aurangabad | 2 | 2 | 100 | 5 | 3 | 3 | 100 | 5 | 1 | 1 | 100 | 5 |
| Banka | 9 | 7 | 78 | 4 | 1 | 1 | 100 | 5 | 0 | 0 | 0 | 0 |
| Begusarai | 2 | 2 | 100 | 5 | 0 | 0 | 0 | 0 | 1 | 1 | 100 | 5 |
| Kaimur | 8 | 7 | 88 | 5 | 4 | 4 | 100 | 5 | 0 | 0 | 0 | 0 |
| Bhagalpur | 0 | 0 | 0 | 0 | 0 | 0 | 0 | 0 | 0 | 0 | 0 | 0 |
| Bhojpur | 3 | 3 | 100 | 5 | 1 | 1 | 100 | 5 | 5 | 4 | 80 | 5 |
| Buxar | 3 | 2 | 67 | 4 | 2 | 2 | 100 | 5 | 1 | 1 | 100 | 5 |
| Darbhanga | 5 | 5 | 100 | 5 | 3 | 3 | 100 | 5 | 0 | 0 | 0 | 0 |
| East Champaran | 0 | 0 | 0 | 0 | 0 | 0 | 0 | 0 | 0 | 0 | 0 | 0 |
| Gaya | 5 | 5 | 100 | 5 | 0 | 0 | 0 | 0 | 0 | 0 | 0 | 0 |
| Gopalganj | 13 | 13 | 100 | 5 | 1 | 1 | 100 | 5 | 6 | 6 | 100 | 5 |
| Jamui | 17 | 16 | 94 | 5 | 12 | 12 | 100 | 5 | 3 | 3 | 100 | 5 |
| Jehanabad | 3 | 3 | 100 | 5 | 2 | 2 | 100 | 5 | 5 | 5 | 100 | 5 |
| Katihar | 10 | 10 | 100 | 5 | 7 | 7 | 100 | 5 | 1 | 1 | 100 | 5 |
| Khagaria | 1 | 1 | 100 | 5 | 0 | 0 | 0 | 0 | 0 | 0 | 0 | 0 |
| Kishanganj | 0 | 0 | 0 | 0 | 0 | 0 | 0 | 0 | 0 | 0 | 0 | 0 |
| Lakhisarai | 0 | 0 | 0 | 0 | 0 | 0 | 0 | 0 | 0 | 0 | 0 | 0 |
| Madhepura | 0 | 0 | 0 | 0 | 0 | 0 | 0 | 0 | 2 | 2 | 100 | 5 |
| Madhubani | 25 | 25 | 100 | 5 | 15 | 15 | 100 | 5 | 22 | 22 | 100 | 5 |
| Munger | 13 | 13 | 100 | 5 | 2 | 2 | 100 | 5 | 4 | 4 | 100 | 5 |
| Muzaffarpur | 14 | 13 | 93 | 5 | 16 | 15 | 94 | 5 | 29 | 29 | 100 | 5 |
| Nalanda | 12 | 12 | 100 | 5 | 14 | 13 | 93 | 5 | 10 | 10 | 100 | 5 |
| Nawada | 11 | 10 | 91 | 5 | 15 | 15 | 100 | 5 | 33 | 33 | 100 | 5 |
| Patna | 28 | 27 | 96 | 5 | 11 | 11 | 100 | 5 | 5 | 5 | 100 | 5 |
| Purnia | 2 | 2 | 100 | 5 | 2 | 2 | 100 | 5 | 6 | 6 | 100 | 5 |
| Rohtas | 15 | 15 | 100 | 5 | 3 | 3 | 100 | 5 | 1 | 1 | 100 | 5 |
| Saharsa | 4 | 4 | 100 | 5 | 4 | 4 | 100 | 5 | 1 | 1 | 100 | 5 |
| Samastipur | 8 | 8 | 100 | 5 | 3 | 3 | 100 | 5 | 6 | 6 | 100 | 5 |
| Saran | 2 | 2 | 100 | 5 | 1 | 1 | 100 | 5 | 2 | 2 | 100 | 5 |
| Sheikhpura | 1 | 1 | 100 | 5 | 4 | 4 | 100 | 5 | 1 | 1 | 100 | 5 |
| Sheohar | 1 | 0 | 0 | 0 | 1 | 0 | 0 | 0 | 2 | 2 | 100 | 5 |
| Sitamarhi | 3 | 3 | 100 | 5 | 0 | 0 | 0 | 0 | 1 | 1 | 100 | 5 |
| Siwan | 3 | 2 | 67 | 4 | 0 | 0 | 0 | 0 | 0 | 0 | 0 | 0 |
| Supaul | 5 | 5 | 100 | 5 | 2 | 2 | 100 | 5 | 6 | 6 | 100 | 5 |
| Vaishali | 11 | 10 | 91 | 5 | 4 | 4 | 100 | 5 | 8 | 8 | 100 | 5 |
| West Champaran | 2 | 2 | 100 | 5 | 2 | 2 | 100 | 5 | 7 | 7 | 100 | 5 |
| **Total** | **246** | **233** | **95** | **-** | **142** | **139** | **98** | **-** | **171** | **170** | **99.4** | **-** |

### Flexibility

Flexibility was assessed as the proportion of outbreaks reported for diseases outside the IDSP surveillance list. In 2016, outbreaks of conditions outside the IDSP list were not reported from any district. In 2017, one district (2.6%) reported such an outbreak - Sheikhpura reported Kala azar. In 2018, two districts (5.3%) - Nawada and Purnia each reported one outbreak of Kala azar. The flexibility score was 0 for all districts in 2016; one district (2.6%) scored above 0 in 2017 and two districts (5.3%) in 2018.

### Expert opinion and weightage factors

Twenty-five public health experts participated in the expert opinion survey (Table 2). The group comprised 19 males and 6 females with qualifications ranging from MBBS to PhD and experience in communicable disease surveillance ranging from less than one year to 34 years. The median weightage score assigned to each attribute was: Timeliness - 5, Representativeness - 4, Acceptability - 5, Completeness - 5, Relative Sensitivity - 4 and Flexibility - 4. The maximum possible composite raw score was 135. All district composite scores were scaled proportionally to 100.

### Composite performance scores

District-wise composite performance scores for 2016, 2017 and 2018 are presented in Table 7. The mean composite score across all 38 districts was 26.97 (SD 12.95) in 2016, 23.47 (SD 16.75) in 2017 and 22.61 (SD 15.45) in 2018. All three mean scores fell within a narrow range of 23–27 out of 100, indicating consistently low overall performance across the study period. The increasing standard deviation over time reflects widening disparity in performance between districts.

**Table 7:**
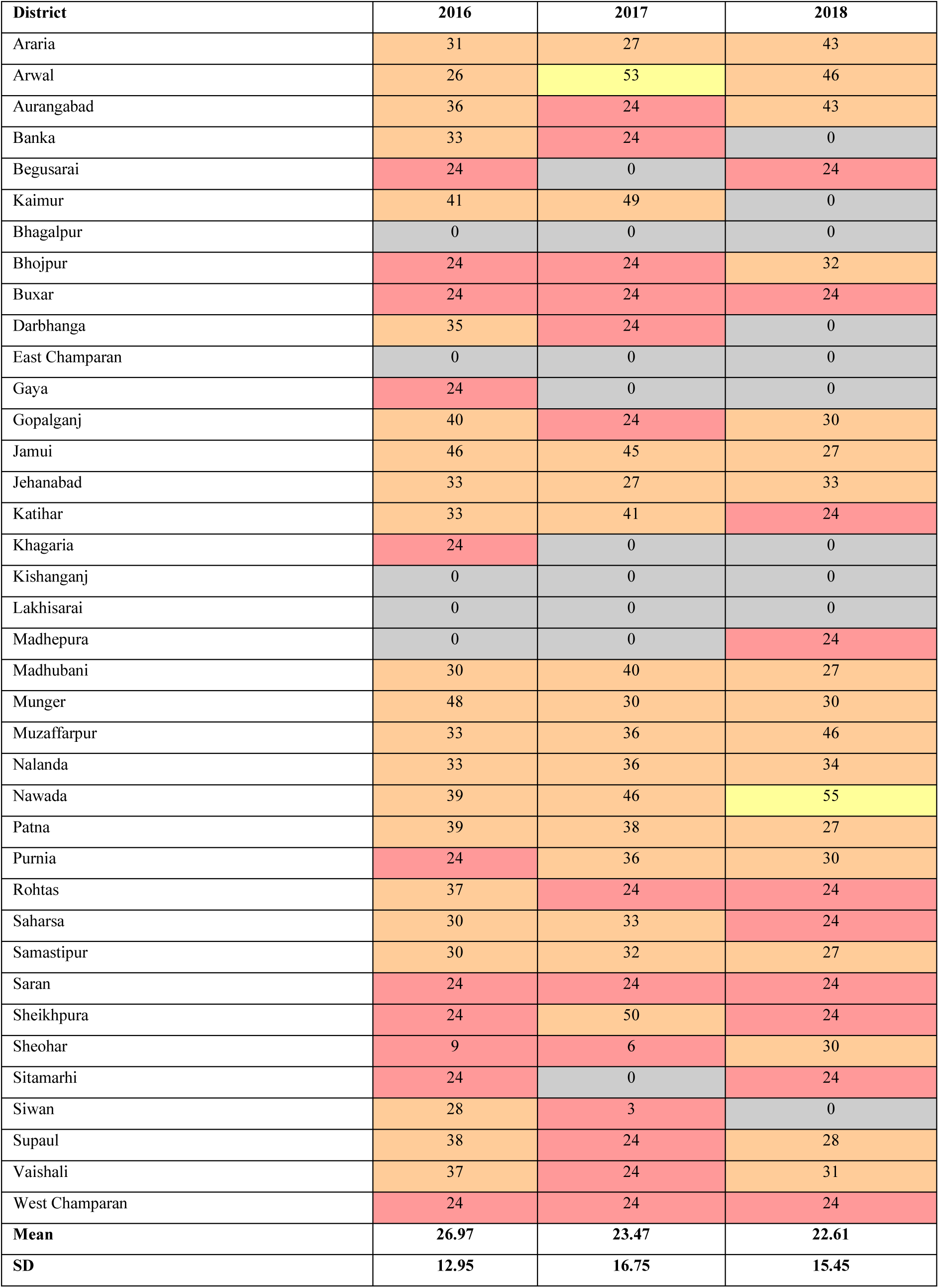
District-wise Composite Performance Evaluation Scores, Bihar, 2016–2018 (Maximum Score: 100)

In 2016, four districts (10.5%) scored 40 or above — Munger (48), Jamui (46), Kaimur (41) and Gopalganj (40); five districts (13.2%) scored 0. In 2017, seven districts (18.4%) scored 40 or above —Arwal (53), Sheikhpura (50), Kaimur (49), Nawada (46), Jamui (45), Katihar (41) and Madhubani (40); nine districts (23.7%) scored 0. In 2018, five districts (13.2%) scored 40 or above — Nawada (55), Muzaffarpur (46), Arwal (46), Aurangabad (43) and Araria (43); ten districts (26.3%) scored 0.

The highest composite score recorded in each year improved progressively — 48 in 2016, 53 in 2017 and 55 in 2018. Districts were ranked annually based on their composite performance score as presented in Table 7. Four districts (10.5%) - Bhagalpur, East Champaran, Kishanganj and Lakhisarai — scored 0 in all three years.

The geographic distribution of composite performance scores across Bihar for 2016, 2017 and 2018 is presented in Figure 1. Districts in the eastern and northeastern zones — including Bhagalpur, Kishanganj and East Champaran — showed consistently low scores across all three years. Districts in the central and southern zones demonstrated greater variability over time. Nawada, in the south, showed the most notable improvement, rising from a mid-range score in 2016 to the highest composite score in 2018.

## DISCUSSION

This study assessed the quality of IDSP outbreak detection and response in Bihar across six surveillance attributes and developed a weighted composite performance evaluation scoring framework using routine program data. The framework was applied at district level over three consecutive years, producing comparable and trackable performance scores across all 38 districts. To the best of our knowledge, this is the first study to develop a dynamic, routine-data-based composite performance scoring framework for IDSP outbreak detection and response [30, 34].

### Surveillance quality attributes

Timeliness was the poorest-performing attribute across all three years. The proportion of outbreaks notified within 48 hours of onset remained below 15% in all three years — 4.9% in 2016, 12.0% in 2017 and 5.8% in 2018. Timeliness received the highest expert weightage (median score 5), reflecting its recognised primacy in outbreak control [1, 36]. Poor timeliness in IDSP has been reported from Andhra Pradesh [26] and Uttarakhand [23]. A review of core IDSP indicators over 2009–2019 reported timeliness peaking at 74% in 2013 and declining to 41% by 2018 [37]. The use of mobile-based reporting demonstrated timeliness gains of 12–30% in Andhra Pradesh [38], suggesting digital platforms offer a practical route to improvement.

Private healthcare providers did not report a single outbreak in any district across all three years. The acceptability score was 0 for all 38 districts across the entire study period. The expert panel assigned acceptability the highest weightage alongside timeliness, reflecting its recognised programmatic importance. Similar findings of absent or negligible private sector participation in IDSP have been reported from Andhra Pradesh [29] and North 24 Parganas, West Bengal [39]. The principal barriers to private sector reporting include limited awareness of surveillance obligations, absence of structured reporting mechanisms and competing clinical workload [40]. Intersectoral coordination and structured engagement with private providers remain unaddressed gaps in IDSP.

Completeness was the strongest-performing attribute. Over 95% of reported outbreaks carried Time, Place and Person information across all three years. This finding reflects adequate capture of outbreak information within the IDSP network once an outbreak is reported. The increasing number of districts not reporting any outbreak – 6 (15.8%) in 2016 rising to 11 (28.9%) in 2017 - indicates gaps in the reach of the reporting network rather than deficiencies in the quality of reports received. Similar findings of high completeness have been reported in national and state-level IDSP evaluations [37, 39].

Representativeness declined over the study period, with the proportion of blocks reporting outbreaks falling from 26.2% in 2016 to 16.7% in 2018. The number of districts not meeting the representativeness criterion increased from 5 (13.2%) to 10 (26.3%). Similar patterns of peripheral underreporting have been documented in integrated surveillance systems in Ghana [41] and Ethiopia [42]. For relative sensitivity, the reference district shifted across years — Munger in 2016, Arwal in 2017 and Nawada in 2018. The use of the highest-reporting district as a proxy reference provides a practical comparative measure in the absence of a gold standard [3]. The number of districts not meeting any relative sensitivity criterion increased from 5 (13.2%) in 2016 to 10 (26.3%) in both 2017 and 2018.

The detection of outbreaks caused by Kala azar – a condition outside the IDSP surveillance list managed under a dedicated vertical elimination program - in 2017 and 2018 demonstrates a degree of system flexibility. Diseases currently managed under vertical programs, including tuberculosis and acute encephalitis syndrome, may benefit from integration within IDSP through this flexibility.

### Composite performance scoring framework

The composite scoring framework produced a single, comparable and trackable measure of overall district performance that individual attribute scores assessed separately cannot provide. The mean composite score remained in a narrow range of 23–27 out of 100 across three years, indicating consistently low overall performance. A finding emerging only from the composite analysis is the widening inter-district disparity — the standard deviation increased from 12.95 in 2016 to 16.75 in 2017. The highest composite score improved progressively from 48 to 55 across the three years, while the number of districts scoring 0 increased from 5 (13.2%) to 10 (26.3%). Four districts (10.5%) scored 0 in all three years, indicating a structural surveillance gap that the framework identifies and can track over time. Such patterns are not identifiable from individual attribute analyses.

The composite score produced for each district enables ranking, which can serve as an evidence base for periodic program reviews, targeted supportive supervision and priority resource allocation to consistently low-performing districts.

The framework design is deliberately modular and configurable. Indicators can be added as data availability improves or removed where routine data do not capture them. Scoring cut-offs can be adjusted to reflect program maturity. Indicator scoring direction is configurable depending on programmatic context, as described in the acceptability indicator. Expert-derived weightages can be re-derived locally as program priorities evolve. The framework demonstrated here at district level can equally be configured at block level, enabling district managers to grade the performance of their own administrative blocks and identify specific geographic areas for targeted intervention. The same framework scales upward to state and national level without modification.

The framework was applied on an annual basis in this study. It can be configured for any time period — monthly, quarterly or any interval aligned with the program’s review cycle — making it adaptable to different monitoring frequencies without any structural modification.

The geographic visualisation of composite scores, as demonstrated in Figure 1, illustrates how this framework can function as a performance monitoring dashboard. Four performance categories are applied — Red (0–25), Light Red (25–50), Yellow (50–75) and Green (75–100) — providing an instantly interpretable picture of district performance. The number and boundaries of categories are themselves configurable: program managers may choose two categories (below and above a defined threshold) or any number appropriate to their decision-making context. Digital health information platforms that capture routine surveillance data provide the infrastructure within which such a dashboard can function on a continuous, near-real-time basis — including India’s Integrated Health Information Platform [43].

Similar digital surveillance initiatives at state level, such as the Unified Disease Surveillance Program (UDSP) of Uttar Pradesh [44], further demonstrate the enabling infrastructure within which such a framework can be operationalised.

India’s Vision 2035: Public Health Surveillance endorses this direction [35], and this study responds directly to the Joint Monitoring Mission 2015 recommendation for a self-assessment mechanism for IDSP [32].

### Limitations

The composite scoring framework has not been validated against an independent measure of surveillance performance. The relationship between composite scores and actual outbreak detection capacity remains to be established. Expert weightages were derived from a single-round opinion survey of 25 participants — a formal Delphi process would provide more robust and stable weightages. The indicator set was constrained by data available in routine weekly outbreak reports — attributes recognised in established evaluation frameworks, including positive predictive value, stability and cost, could not be assessed [1, 21]. Routine data quality determines score accuracy — errors or omissions in source data are reflected directly in attribute scores. The relative sensitivity indicator uses the highest-reporting district as a proxy reference, providing a comparative measure but not true sensitivity of the surveillance system [3].

Acknowledged limitations of routine surveillance data — including incomplete reporting, fragmentation and validity concerns — do not diminish the utility of this framework. Rather, such limitations are themselves captured and quantified through the attribute scoring process, making data quality problems visible and actionable. The framework operates on the principle that systematic utilisation of available program data for decision-making and mid-course correction is preferable to deferring action pending ideal data conditions.

## CONCLUSION

IDSP generates continuous outbreak data through weekly reports that remain underutilised for system performance evaluation. This study demonstrates that routine program data, systematically analysed across six surveillance quality attributes, can produce a meaningful composite performance score for every district without any additional data collection. The framework was applied across all 38 districts of Bihar over three consecutive years, revealing consistently low overall performance, entirely absent private sector participation, and widening inter-district disparity.

The framework is modular, bidirectionally configurable in indicator scoring direction, and scalable across administrative levels — from block to national. Scoring cut-offs, performance categories and weightage factors are all adjustable to program context and maturity. Geographic visualisation of composite scores demonstrates the dashboard potential of this approach, translating complex multi-attribute data into an instantly interpretable map for program managers.

This framework is not intended as a replacement for commissioned evaluations, which provide comprehensive long-term programmatic assessment, or for academic research, which addresses specific hypotheses through rigorous methodology. It is designed to complement both by providing a simple, program-embedded tool for ongoing performance monitoring and mid-course correction at the operational level.

Future research should focus on validation of the composite score against independent measures of surveillance performance, re-derivation of weightages through a formal Delphi process, and integration of the framework within digital health information platforms to enable real-time, continuous performance monitoring of IDSP at scale across all states.

## Data Availability

All data produced in the present study are available upon reasonable request to the authors

## REFERENCES

1. Centers for Disease Control and Prevention. Updated guidelines for evaluating public health surveillance systems: recommendations from the guidelines working group. MMWR Recomm Rep. 2001;50(RR-13):1–35.

2. World Health Organization. An integrated approach to communicable disease surveillance. Epidemiological Bulletin PAHO. 2000;13(1):1–16.

3. Centers for Disease Control and Prevention. Framework for Evaluating Public Health Surveillance Systems for Early Detection of Outbreaks: Recommendations from the CDC Working Group. MMWR Recomm Rep. 2004;53(RR05):1–11.

4. Ministry of Health and Family Welfare, Government of India. Integrated Disease Surveillance Project: Training Manual for Paramedical Staff for Hospital Based Disease Surveillance. New Delhi: MoHFW; 2004.

5. M’ikanatha N, Lynfield R, Julian K, Van Beneden C, De Valk H. Infectious Disease Surveillance. Malden MA: Wiley-Blackwell; 2007.

6. Franco LM, Setzer J, Banke KK. Improving performance of IDSR at district and facility levels: Experiences in Tanzania and Ghana in making IDSR operational. 2006.

7. Lukwago L, Nanyunja M, Ndayimirije N, et al. The implementation of Integrated Disease Surveillance and Response in Uganda: a review of progress and challenges between 2001 and 2007. Health Policy Plan. 2012;28(1):30–40.

8. Nsubuga P, Eseko N, Tadesse W, et al. Structure and performance of infectious disease surveillance and response, United Republic of Tanzania, 1998. Bull World Health Organ. 2002;80(3):196–203.

9. Calain P. From the field side of the binoculars: a different view on global public health surveillance. Health Policy Plan. 2007;22(1):13–20.

10. World Health Organization Regional Office for Africa. Technical guidelines for integrated disease surveillance and response in the African region. Harare: WHO AFRO; 2001.

11. World Health Organization Regional Office for South East Asia. Regional strategic plan for integrated disease surveillance 2002–2010. New Delhi: WHO SEARO; 2003.

12. Kasolo F, Roungou JB, Perry H. Technical Guidelines for Integrated Disease Surveillance and Response in the African Region. 2nd ed. Atlanta/Brazzaville: CDC/WHO; 2010.

13. Ministry of Health and Family Welfare, Government of India. Integrated Disease Surveillance Project: Project Implementation Plan. New Delhi: MoHFW; 2004. Available from: http://www.idsp.nic.in/

14. Government of India. Integrated Disease Surveillance Project: Operations manual for district surveillance unit. New Delhi: National Institute for Communicable Diseases, MoHFW; 2002.

15. European Centre for Disease Prevention and Control. Data quality monitoring and surveillance system evaluation — A handbook of methods and applications. Stockholm: ECDC; 2014.

16. World Health Organization. Protocol for the assessment of national communicable disease surveillance and response systems: Guidelines for assessment teams. Geneva: WHO; 2001.

17. World Health Organization. Communicable disease surveillance and response system: A guide to monitoring and evaluation. Lyon: WHO; 2006.

18. World Health Organization. Overview of the WHO framework for monitoring and evaluating surveillance and response systems for communicable diseases. Wkly Epidemiol Rec. 2004;36:322–6.

19. Centers for Disease Control and Prevention. Guidelines for evaluating surveillance systems. MMWR Suppl. 1988;37(S-5):1–18.

20. Drewe JA, Hoinville LJ, Cook AJ, Floyd T, Stärk KD. Evaluation of animal and public health surveillance systems: a systematic review. Epidemiol Infect. 2012;140(4):575–90.

21. Calba C, Goutard FL, Hoinville L, et al. Surveillance systems evaluation: a systematic review of the existing approaches. BMC Public Health. 2015;15:448.

22. Watkins RE, Eagleson S, Hall RG, Dailey L, Plant AJ. Approaches to the evaluation of outbreak detection methods. BMC Public Health. 2006;6:263.

23. Srivastava DK, Venkatesh S, Pandey S, Shankar R, Pillai DS. Completeness and timeliness of reporting under integrated disease surveillance project (IDSP) in rural surveillance unit of Nainital district of Uttarakhand, India. Indian J Prev Soc Med. 2009;40(3–4).

24. Kumar A, Goel MK, Jain RB, Khanna P. Tracking the Implementation to Identify Gaps in Integrated Disease Surveillance Program in a Block of District Jhajjar (Haryana). J Family Med Prim Care. 2014;3(3):213–5.

25. Borker S, Venugopalan PP. Evaluation of the Integrated Disease Surveillance Project Training at Kannur district of North Kerala. Indian J Public Health. 2010;54(1).

26. Singh V. Importance of Timeliness, Completeness and Timely Response to an Outbreak in disease surveillance: An evaluation of the critical components of IDSP in Andhra Pradesh, India. Research Fellowship under the PHFI-UKC Wellcome Trust Capacity Building Programme; 2010–2012.

27. Gupta SN, Gupta N, Gupta S. Evaluation of Diarrhoeal Diseases Surveillance System of District Kangra, Himachal Pradesh, India, 2007. J Res Med Edu Ethics. 2013;3(1).

28. Dan A, Roy B, De KK, Pasi AR, Jalaluddeen M. Evaluation of system for surveillance of dengue in Hooghly district of West Bengal — India. Asian Academic Research Journal of Multidisciplinary. 2016;3(3).

29. Phalkey RK, Shukla S, Shardul S, et al. Assessment of the core and support functions of the Integrated Disease Surveillance system in Maharashtra, India. BMC Public Health. 2013;13:575.

30. Singh V, Mohan J, Rao UP, Dandona L, Heymann D. An Evaluation of the Key Indicator Based Surveillance System for International Health Regulations (IHR)-2005 Core Capacity Requirements in India. Online J Public Health Inform. 2014;6(1):e121.

31. Singh V, Mohan J, Rao UP, Dandona L, Heymann D. Validity of the Surveillance Quality Indicators — Timeliness and Completeness — in Surveillance Systems with Variable Data Quality. Online J Public Health Inform. 2014;6(1):e176.

32. Joint Monitoring Mission. Integrated Disease Surveillance Programme 2015. New Delhi: Directorate General of Health Services, Ministry of Health and Family Welfare, Government of India; 2015.

33. McNabb SJN, Chungong S, Ryan M, et al. Conceptual framework of public health surveillance and action and its application in health sector reform. BMC Public Health. 2002;2:2.

34. Cavallaro KF, Sandhu HS, Hyde TB, Johnson BW, Fischer M, Mayer LW, Clark TA, Pallansch MA, Yin Z, Zuo S, Hadler SC, Diorditsa S, Hasan AS, Bose AS, Dietz V; AMES Study Group. Expansion of syndromic vaccine preventable disease surveillance to include bacterial meningitis and Japanese encephalitis: evaluation of adapting polio and measles laboratory networks in Bangladesh, China and India, 2007–2008. Vaccine. 2015;33(9):1168–75.

35. NITI Aayog. Vision 2035: Public Health Surveillance in India. New Delhi: Government of India; 2020. Available from: https://niti.gov.in/sites/default/files/2020-12/PHS_13_dec_web.pdf

36. Jajosky RA, Groseclose SL. Evaluation of reporting timeliness of public health surveillance systems for infectious diseases. BMC Public Health. 2004;4:29.

37. Mahapatra S, Mishra R, Paul S, Kumari S, Jha R. Integrated Disease Surveillance Program (IDSP) implementation in Bihar since inception in 2009: A critical review. Working Paper 04/Version 01. Patna: The Centre for Health Policy, Asian Development Research Institute (ADRI); March 2020. Available from: https://www.adriindia.org/images/paper/1603351542Book042020_Ok.pdf

38. Singh V, Madhusudana Rao B, Mohan J. An evaluation of mobile phone technology use for Integrated Disease Surveillance Project (IDSP) in Andhra Pradesh, India. Emerg Health Threats J. 2011;4(0).

39. Debnath F, Ponnaiah M. Improved timeliness for reporting of acute diarrhoeal disease under surveillance overtime: Evaluation of integrated disease surveillance programme in North 24 Parganas, West Bengal, India, 2015. Clin Epidemiol Glob Health. 2018;6(4):163–7.

40. Shinde RR, Kembhavi RS, Kuwatada JS, Khandednath TS. To develop a public private partnership model of disease notification as a part of integrated disease surveillance project (IDSP) for private medical practitioners in Mumbai City, India. Glob J Med Public Health. 2012;1(6):1–11.

41. Adokiya MN, Awoonor-Williams JK, Beiersmann C, Müller O. The integrated disease surveillance and response system in northern Ghana: challenges to the core and support functions. BMC Health Serv Res. 2015;15:288.

42. Alemu T, Gutema H, Legesse S, et al. Evaluation of public health surveillance system performance in Dangila district, Northwest Ethiopia. BMC Public Health. 2019;19:1343.

43. World Health Organization India. Next-gen digital platform launched pan India to accelerate outbreak response. New Delhi: WHO India; 2021. Available from: https://www.who.int/india/news-room/detail/14-04-2021-next-gen-digital-platform-launched-pan-india-to-accelerate-outbreak-response

44. Department of Medical Health & Family Welfare, Government of Uttar Pradesh; UP-TSU; IHAT. Unified Disease Surveillance Platform (UDSP): Journey from Idea to Implementation. Lucknow: Government of Uttar Pradesh; 2025. Available from: https://www.ihat.in/wp-content/uploads/2025/03/UDSP.pdf

